# Predicting Mobile Health Clinic Utilization for COVID-19 Vaccination in South Carolina: A Statistical Framework for Strategic Resource Allocation

**DOI:** 10.1101/2024.09.27.24314475

**Authors:** Fatih Gezer, Kerry A. Howard, Kevin J. Bennett, Alain H. Litwin, Kerry K. Sease, Lior Rennert

## Abstract

**Background:** Mobile health clinics (MHCs) are effective tools for providing health services to disadvantaged populations, especially during health emergencies. However, patient utilization of MHC services varies substantially. Strategies to increase utilization are therefore needed to maximize the effectiveness of MHC services by serving more patients in need. The purpose of this study is to develop a statistical framework to identify and prioritize high-risk communities for delivery of MHCs during health emergencies.

**Methods:** Prisma Health MHCs delivered COVID-19 vaccines to communities throughout South Carolina between February 20, 2021, and February 17, 2022. In this retrospective study, we use generalized linear mixed effects model and ordinal logistic regression model to identify factors associated with, and predict, MHC utilization for COVID-19 vaccination by census tract.

**Results:** The MHCs conducted 260 visits to 149 sites and 107 census tracts. The site-level analysis showed that visits to schools (RR=2.17, 95% CI=1.47-3.21), weekend visits (RR=1.38, 95% CI=1.03-1.83), and visits when the resources were limited (term 1: 7.11, 95% CI=4.43-11.43) and (term 2: 2.40, 95% CI=1.76-3.26) were associated with greater MHC utilization for COVID-19 vaccination. MHC placement near existing vaccination centers (RR=0.79, 95% CI=0.68-0.93) and hospitals (RR=0.83, 95% CI=0.71-0.96) decreased utilization. Predictive models identified 1,227 (94.7%) census tracts with more than 250 individuals per MHC visit when vaccine resources were limited. Predictions showed satisfactory accuracy (72.6%). The census tracts with potential of high MHC demand had higher adolescent, 30-44 years old, non-White populations, lower Primary Care Practitioners per 1,000 residents, fewer hospitals, and higher cumulative COVID-19 emergency department visits and deaths (compared to census tracts in the low MHC demand category). After the vaccines became widely available, the demand at MHCs declined.

**Conclusion:** These study findings can be used to improve MHC allocation by identifying and prioritizing medically underserved communities for strategic delivery of these limited resources.

## Introduction

The COVID-19 pandemic has affected the lives of millions worldwide, including nearly 1.2 million deaths in the United States (US).[1] The pandemic has also exacerbated inequalities in health outcomes, compounded with ethnic minority groups and rural communities experiencing less access for testing, vaccination, and treatment services, and greater death rates.[2–8] These consequences underscored the need for strategies to reach vulnerable communities more effectively.[9,10] Mobile health clinics (MHCs) deliver quality healthcare services to medically underserved communities who lack access to healthcare resources and facilities, especially during health emergencies.[11–14] MHCs were used during the COVID-19 pandemic for vaccination in the U.S. and different countries,[15–19] and mainly benefitted by rural communities and medically underserved populations.[11,12,20,21]

However, the inability to effectively identify and prioritize high-risk communities has posed daunting challenges for decision makers and has led to less-than-optimal allocation strategies. This is especially problematic during phases of pandemics when resources are limited, as these phases correlate with periods of high transmission, morbidity, and mortality.[22–24] Less efficient allocation strategies have a disproportionate impact on medically underserved communities. For example, age-based allocation of COVID-19 vaccines adopted by states nationwide lead to inequity in vaccination uptake, with lower rates in economically disadvantaged neighborhoods that were at an increased risk for severe SARS-CoV-2 infection and death.[25–27] Alternatively, one study showed that the inclusion of geographic region into the prioritization process would have led to an estimated 18% decrease in COVID-19 related hospitalizations.[25]

Data-driven approaches can improve emergency planning and overall health outcomes by guiding timely delivery of essential resources to high-risk communities.[28–31] One study showed that prioritization of COVID-19 testing to high-risk areas is twice as likely to detect positive cases compared to random allocation of tests (e.g., based on population sized).[32] Moreover, utilization of MHC services can substantially vary by site location.[33,34] Site visits with low utilization are a missed opportunity to provide health care to individuals in need, and drastically reduce the potential impact of MHC services. Therefore, projecting low- and high-demand areas at granular geographic levels can assist in optimizing the effectiveness of MHC services.[35]

Various studies investigated the characteristics of individuals who used MHCs and community-level factors associated with MHC utilization.[36,37] Individuals who utilized the MHCs tend to be mostly ethnic minorities and uninsured persons.[34,38] MHCs had a higher uptake when they were located at places where the proportions of uninsured and non-white populations were higher, and primary care practitioner rates were lower.[34] Several studies built predictive models for COVID-19 vaccine uptake using structural equation models, machine learning-based approaches, and conceptual models.[39–42] However, these studies were conducted based on data for general populations rather than MHC users hence they did not target medically underserved populations. For such populations, geographic granularity is needed to effectively inform public health interventions, including MHCs.

In this retrospective study, we develop predictive models to predict MHC utilization for COVID-19 vaccination in South Carolina (SC) during different phases of the pandemic. The projection of MHC utilization allows us to signify the low- and high-demand census tracts for MHC utilization and understand their characteristic differences. We also explore MHC logistical factors and community determinants that contribute to greater utilization after the high-demand regions are detected, ultimately aiming to inform policy, improve public health outcomes, and optimally allocate MHCs for greater utilization.

## Methods

### Setting

Prisma Health is a SC-based healthcare organization that serves 1.2 million patients per year.[43] Prisma Health deployed MHCs to increase COVID-19 vaccination in underserved communities between February 20, 2021, and February 17, 2022. A detailed explanation of Prisma Health’s MHC activities for COVID-19 vaccination program is provided in the literature.[34]

### Variables

Data obtained from MHC visits included the name, time, date, and location of the visit site, site type, duration of the visit, and the number of individuals who received COVID-19 vaccines. Site types include churches; public K-12 schools; universities; corporate locations such as business centers, homeless shelters; and other kinds of places such as community and wellness centers, supermarkets, and parks. Based on the data, we categorized the timing of the week (Monday to Thursday, Friday, or Weekend), determined time of the day (morning: before 12 pm, afternoon: from 12 to 4 pm, and evening: after 4 pm), visit number (first, second, and third or more), and zip code and census tract code of the visited sites.

The community-level variables included census tract and zip code level demographic, socioeconomic, and health-related factors. Age, sex, race, ethnicity, median income, unemployment rate, labor force participation, and social vulnerability index (SVI) variables were at the census tract level and linked to census tracts of the MHC site locations. These demographic and socioeconomic variables were obtained from the United States Census Bureau American Community Survey for 2021.[44] SVI is a measure that assesses the resilience of communities when faced with external stresses and is developed by the Agency for Toxic Substances and Disease Registry (ATSDR) at the Centers for Disease Control and Prevention (CDC).[45] The four components of SVI are socioeconomic status, household composition and disability, minority status and language, and housing and transportation.[46] Data related to health care access were obtained from The South Carolina Center for Rural and Primary Healthcare (SCCRPH) and is available at the zip code level.[47] These variables included the number of hospitals, primary care physicians (PCP) per 1000 residents, all-cause mortality rate per 1000 residents, the percentage of uninsured individuals in each zip code, and the percentage of rural areas in each zip code is provided. We also included the number of vaccination centers within a 2-mile radius and the number of hospitals within a 3-mile radius of each MHC location.[48,49] Data for emergency department (ED) COVID-19 hospitalizations and deaths were obtained from the South Carolina Revenue and Fiscal Affairs Office.[50]

### Statistical analysis

We used negative binomial generalized linear mixed effects models to assess the relationship between site-related factors and MHC utilization for COVID-19 vaccination. For the projection of MHC utilization, we aimed a ranking based approach of possible MHC utilization at census tract level. Therefore, we used ordinal logistic regression models for the ordered categorical outcome of the MHC utilization using demographic, socioeconomic, and health-related predictors. Details about the model descriptions are provided in the Supplementary Material.

The ordinal grouping of the MHC utilization consisted of number of individuals within 0-19, 20-49, 50-99, 100-249, 250-399, and more than 400. These groupings were used both for the validation of the models built on available MHC data, and for the projection of MHC utilization for the remaining census tracts in SC. The validation stage is performed by randomly sampling training and validation sets from the entire MHC data available for 106 census tracts. The validation set included 25 randomly sampled census tracts and the remaining data is used for the training of the models. Once the models are trained, we predicted the MHC utilization in 25 census tracts and compared it with the truth. If the predicted category is correctly predicted, we noted is as exact category prediction. However, if the predicted category was different than the truth, we noted the degree of deviation by checking how many categories the prediction was away from the truth, for instance, one group higher than truth or two groups lower than truth. We repeated the random sampling of 25 census tracts for 1,000 times independently, hence we approximated the accuracy of the prediction performance. For the variables available at the ZCTA level, we partitioned the data into census tracts linked to that ZCTA weighting on the census tract population.

Due to significant changes in vaccine eligibility and availability, we stratify the study period before and after March 31, 2021. There were significant policy changes on or near these dates regarding the targeted groups for vaccination in the US. For instance, individuals aged 16 or older become eligible for COVID-19 vaccination on March 31. At the same time, COVID-19 vaccines became more widely available at pharmacies, clinics, and other health-providing organizations.[51] As a sensitivity analysis, we also stratify the time period before and after May 10, 2021, which is the date adolescents aged 12 to 15 were eligible for COVID-19 vaccination.[52]

## Results

### Descriptive summaries

Between February 20, 2021, and February 17, 2022, the Prisma Health MHCs had 260 visits to 149 locations in SC. These visits took place in 59 zip codes (of 424 zip codes) and 107 census tracts (of 1,323 census tracts), and the MHC delivered 12,102 vaccines to 8,545 individuals. Descriptive statistics for the individuals and site types have been provided in previous research.[34] The vaccine uptake at each site visit over one-year period is shown in Figure 1. Box plots are generated based on the site visits in each month represented by the red points. MHCs had higher demand per visit before March 31, 2021, when the vaccination resources were limited and there were restrictions on certain age groups. After this date, although the MHCs increased the frequency of its activities, utilization of MHCs per visit decreased. There was an exception of a school site with visits that occurred in July and August 2021 in which the MHC exceeded 600 vaccinations on both dates.

**Figure 1.**
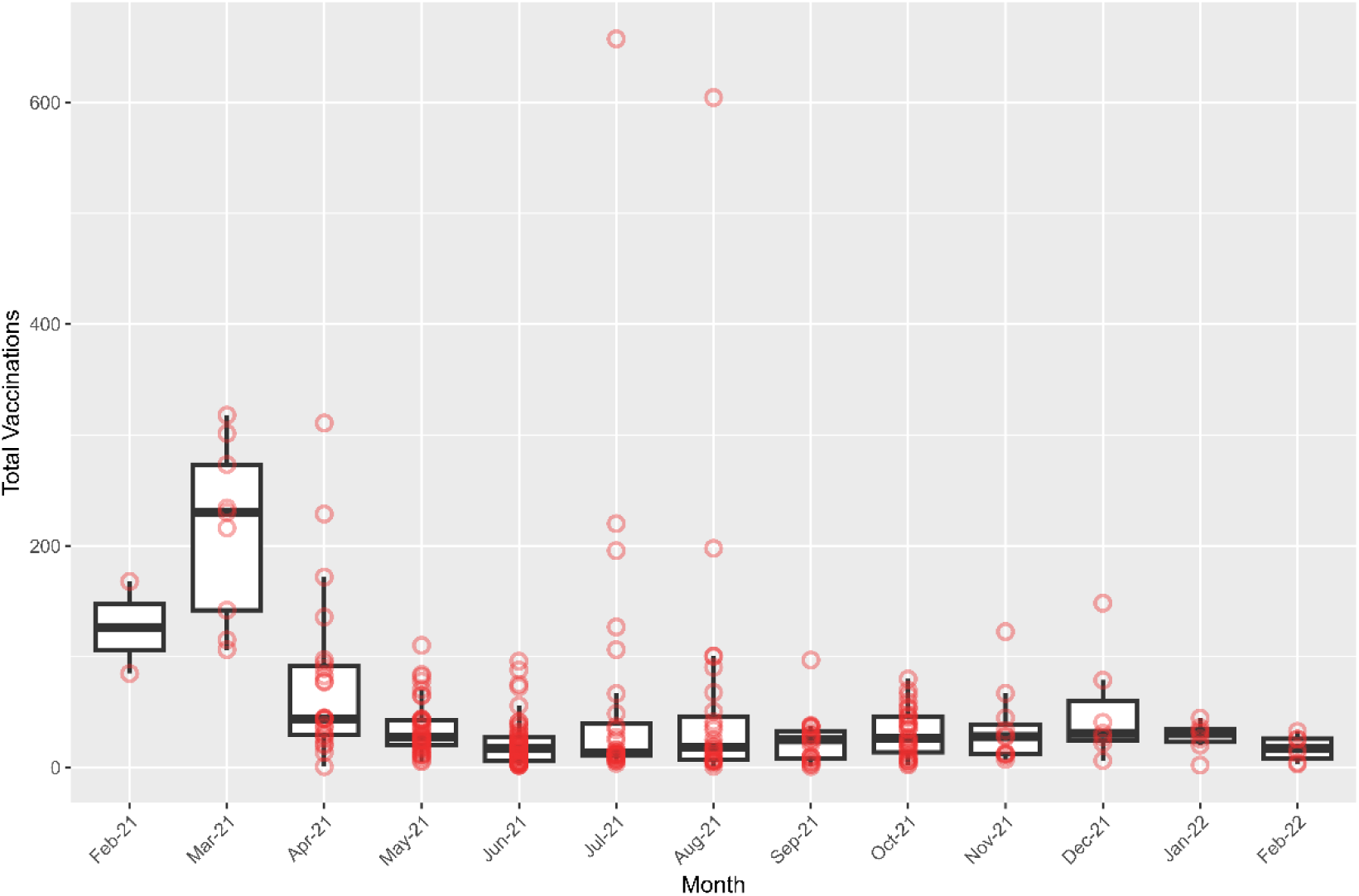
Boxplot of vaccination counts at site visits per month between February 20, 2021, and February 17, 2022. Points show the number of vaccinated individuals at each site visit for different months and constitute the box plot.

Vaccine uptake based on the site type and the day and time of the visit for different vaccination terms (first term: before March 31, 2021, second term: between April 1 and May 9 of 2021, and third term: after May 10, 2021) are summarized in Figure 2. The majority of the MHC events in the first term took place on weekends and mornings at churches, schools, and universities. Although MHCs started visits on different days and times in the second and third terms, overall MHC utilization per visit was not high especially on the third term. Considering all terms, church visits mainly occurred on weekend mornings, and school and corporate visits Monday to Thursday afternoons.

**Figure 2.**
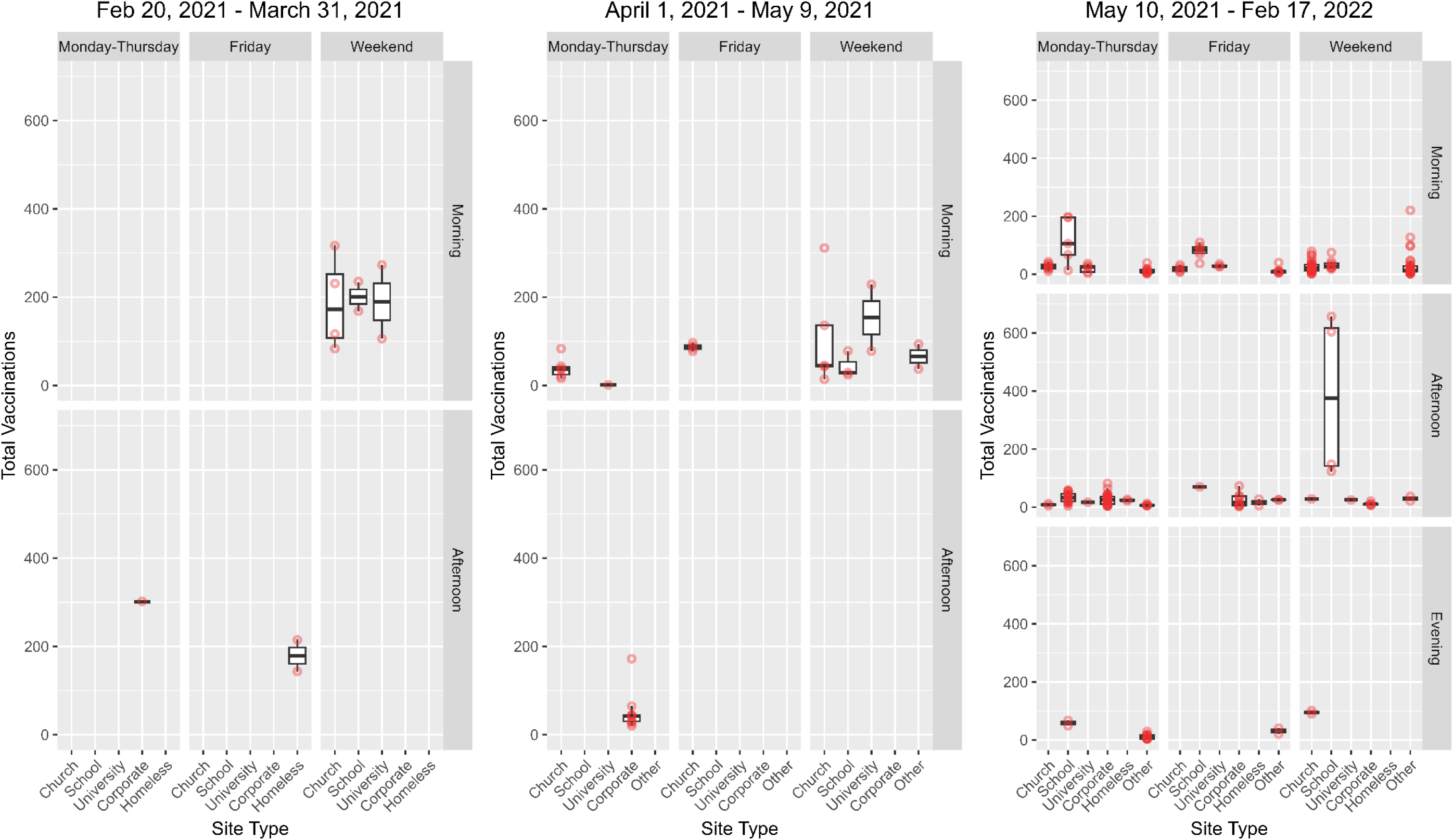
Boxplot of vaccination counts based on different terms (before March 31, 2021, between April 1 and May 9 of 2021, and after May 10, 2021), site types, the day of the week, and the time of the day.

### Site-related factors associated with MHC utilization

The site-related factors associated with MHC utilization are shown in Table 1. Estimated relative risk (RR) with 95% confidence intervals (CI) represents the relative change in the MHC utilization with respect to a standard deviation increase in the continuous predictor variables, and a category change in the categorical variables. School visits and visits on Weekends, and visits within the first term (between February 20, 2021, and March 31, 2021) and second term (between April 1, 2021 and February 17, 2022) were associated with higher MHC utilization. Also, MHCs gained greater utilization at their second visits to a certain site. Having nearby vaccination centers and hospitals, and other types of site visits than schools, churches, corporate locations and homeless shelters decreased MHC utilization.

**Table 1:** Results for negative binomial models that are adjusted for vaccination term, site type, day of the week, time of the day, visit number, visit duration, and population.

|  | RR | CI | P-value |
| --- | --- | --- | --- |
| **Vaccination Term (Ref: Third term)** |  |  |  |
| First Term | 7.11 | (4.43 - 11.43) | **<0.001** |
| Second Term | 2.40 | (1.76 - 3.26) | **<0.001** |
| **Site type (Ref: Church)** |  |  |  |
| School | 2.17 | (1.47 - 3.21) | **<0.001** |
| University | 1.22 | (0.68 - 2.17) | 0.509 |
| Corporate | 1.19 | (0.71 - 1.99) | 0.520 |
| Homeless | 1.17 | (0.49 - 2.80) | 0.731 |
| Other | 0.68 | (0.48 - 0.97) | **0.033** |
| **Day of the week (Ref: Monday – Thursday)** |  |  |  |
| Friday | 1.32 | (0.97 - 1.81) | 0.079 |
| Weekend | 1.38 | (1.03 - 1.83) | **0.029** |
| **Time of the day (Ref: Morning)** |  |  |  |
| Afternoon | 1.17 | (0.78 - 1.74) | 0.449 |
| Evening | 1.44 | (0.84 - 2.47) | 0.184 |
| **Visit number (Ref: First visit)** |  |  |  |
| Second | 1.21 | (1.01 - 1.45) | **0.036** |
| Third or more | 0.96 | (0.75 - 1.25) | 0.778 |
| Visit duration | 1.00 | (0.89 - 1.12) | 0.993 |
| Population | 1.06 | (0.92 - 1.22) | 0.429 |
| Number of nearby vaccination centers | 0.79 | (0.68 - 0.93) | **0.003** |
| Number of nearby hospitals | 0.83 | (0.71 - 0.96) | **0.015** |

### Model Validation

The agreement between the predicted and actual category of MHC utilization is summarized in Table 2 using term-based models (term 1: before March 31, 2021, term 2: after April 1, 2021). Overall, 72.6% of model predictions were within +/- 1 of the true vaccine uptake group as described in the methods. The models predicted the exact category of the outcome in 30.5% of the validation observations, and they predicted one group either a higher or lower category than the actual category for 42.1% of the validation observations.

**Table 2:** Results for prediction accuracy. Number of census tracts that have the same and deviated observed and predicted category for term-based predictions (term 1: before March 31, 2021, term 2: after April 1, 2021).

|  | Term-based Predictions |
| --- | --- |
| Predicted Category | N (%) |
| 5 groups higher than truth | - |
| 4 groups higher than truth | - |
| 3 groups higher than truth | 197 (0.8) |
| 2 groups higher than truth | 2,598 (10.4) |
| **1 group higher than truth** | **5,394 (21.6)** |
| **Exact group** | **7,635 (30.5)** |
| **1 group lower than truth** | **5,136 (20.5)** |
| 2 group lower than truth | 2,974 (11.9) |
| 3 group lower than truth | 926 (3.7) |
| 4 group lower than truth | 140 (0.6) |
| 5 group lower than truth | - |

### Vaccine projections for census tracts

We projected the MHC utilization for all census tracts of SC. These models use the characteristics of census tracts that MHCs visited as the predictors and projected the ordinal category of MHC utilization in other census tracts. The projections are performed prior and post March 31, 2021. The number of census tracts in each category of predicted MHC utilization is shown in Table 3 for both cases. Projected MHC utilization per visit prior to March 31, 2021, was substantially higher at most census tracts (Figure 3). Results for 12-months and for cutoff term of May 10, 2021, are provided in Figure S1 in the Supplementary Material. There were 32 (2.5%) census tracts that were projected to receive more than 400 visitors at a single MHC visit, and 1,195 (92.2%) census tracts projected to receive relatively high utilization with 250 to 399 individuals, whereas only 69 (5.3%) census tracts were estimated to be utilized by under 249 individuals. On the other hand, the models predicted overall low MHC utilization starting from April 1, 2021, where only 181 (14.0%) census tracts would receive between 50 and 99 individuals, and 86.1% of census tracts would have low utilization.

**Figure 3:**
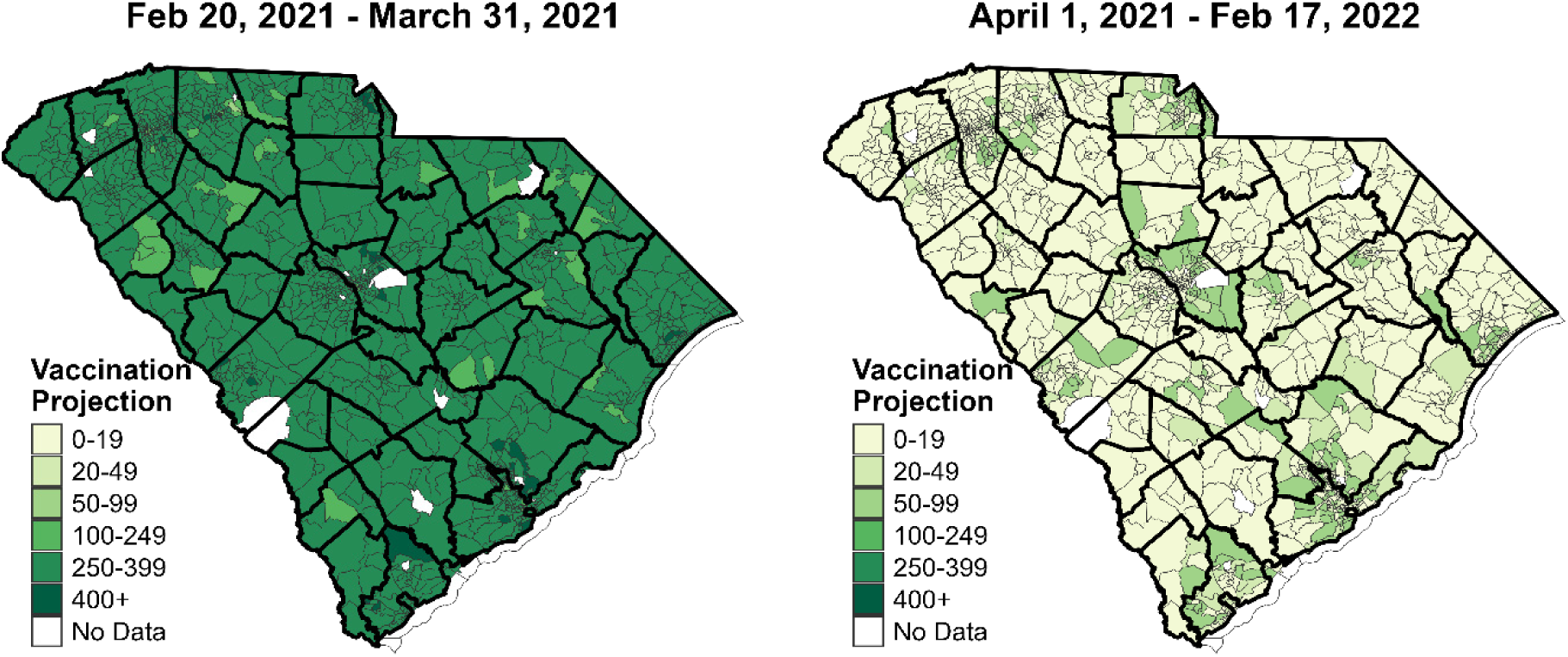
Projected MHC utilization for COVID-19 vaccination at census tracts at different time periods (pre and post March 31, 2021).

**Table 3:** Number of census tracts (N) for each category of the projected MHC utilization. Projections are made for an MHC visit at any time before March 31, 2021, and after April 1, 2021.

| Grouping | Before<br>March 31<br>N = 1,296 (%) | After<br>April 1<br>N = 1,296 (%) |
| --- | --- | --- |
| 0-19 | - | 973 (75.0) |
| 20-49 | - | 142 (11.0) |
| 50-99 | 4 (0.3) | 181 (14.0) |
| 100-249 | 65 (5.0) | - |
| 250-399 | 1,195 (92.2) | - |
| 400+ | 32 (2.5) | - |

Census tracts categorized in the highest and lowest MHC utilization categories had substantial differences in some of their community-level characteristics (Table 4). Compared to the low MHC utilization category, the highest category census tracts had a higher proportion of individuals under 18 years of age and 30-44 age group, a higher proportion of non-White and Hispanic individuals. More importantly, these census tracts had lower PCP rates, lower number of hospitals, higher cumulative COVID-19 ED visits and deaths prior to MHCs were deployed on February 20, 2021. Sensitivity analyses for the cutoff date of May 10, 2021, and for 12-month term are also performed, and the results are provided Tables S2-S6, respectively.

**Table 4:** Characteristics of census tracts that are projected to have high (groups: 250-399 and 400+) and low (groups: 50-99 and 100-249) MHC utilization for COVID-19 vaccination before March 31, 2021. Median values with IQR and p-values for significance of the difference of medians are provided.

|  | High Utilization<br>N = 1,227 | Low Utilization<br>N = 69 | P-value |
| --- | --- | --- | --- |
| % Age under 18 | 21.6 (18.0-25.1) | 20.1 (13.9-23.4) | 0.044 |
| % Age 18-29 | 14.1 (11.0-17.5) | 16.3 (12.6-34.7) | **0.001** |
| % Age 30-44 | 18.3 (15.0-21.5) | 16.2 (11.5-19.7) | **0.011** |
| % Age 45-64 | 26.5 (23.0-29.9) | 25.7 (16.9-31.0) | 0.263 |
| % Age over 65 | 17.4 (13.4-21.8) | 16.7 (11.9-21.7) | 0.398 |
| % Male | 48.5 (45.9-51.1) | 49.8 (46.9-53.0) | **0.030** |
| % Non-white | 35.6 (20.5-55.1) | 25.8 (9.1-37.5) | **0.009** |
| % Hispanic | 4.8 (2.8-7.9) | 3.7 (2.7-5.9) | **0.022** |
| SVI | 0.5 (0.3-0.8) | 0.6 (0.3-0.8) | 0.053 |
| Income (×\$1000) | 55.3 (42.6-71.3) | 46.9 (34.0-62.6) | **0.007** |
| % Unemployed | 4.6 (2.6-7.6) | 4.2 (2.6-6.9) | **<0.001** |
| PCP rate | 0.3 (0.0-0.8) | 3.9 (0.5-16.0) | **<0.001** |
| Hospitals | 0.0 (0.0-1.0) | 0.9 (0.2-1.5) | **<0.001** |
| % Uninsured | 10.9 (8.8-12.3) | 10.2 (7.7-11.6) | 0.426 |
| Mortality rate | 51.3 (40.7-60.7) | 50.3 (38.3-60.3) | 0.797 |
| % in Poverty | 11.0 (7.1-15.7) | 14.0 (10.2-17.3) | **0.006** |
| % in Rural | 28.0 (5.0-60.0) | 15.0 (0.2-62.4) | 0.102 |
| COVID-19 ED visits | 1,189 (658-2,012) | 846 (504-1,260) | **0.002** |
| COVID-19 deaths | 64 (35-99) | 41 (22-70) | **0.002** |

## Discussion

This study aims to serve as a tool to improve the MHCs’ activities by providing important insights from the utilization of MHCs during COVID-19 vaccination across South Carolina, highlighting the potential to optimize resource allocation by identifying high-uptake areas and understanding the factors influencing MHCs’ success. The findings underscore the importance of strategically deploying MHCs to maximize vaccine reach, particularly among underserved and vulnerable populations.

Operational factors including site type (e.g., church, school, etc.), day, and time of the week were significantly associated with vaccine uptake. Notably, visits to schools and visits conducted on weekends were linked to higher vaccine uptake. During the pre- and post-March 31, 2021, 72.7% and 38.6% of MHC visits were conducted on weekends with median utilization of 199 (IQR: 112 - 243) and 23 (IQR: 10 – 38) per visit, respectively.[34] Although the school visits were similar for pre- and post-March 31, 2021 with 18.2% and 18.5%, the utilization per visit was 200 (IQR: 184 - 217) and 45 (IQR: 24 - 77) making highest utilized location for post-March 31, 2021 followed by universities 27 (IQR: 17 - 36). These findings suggest that MHCs can achieve a more significant impact by targeting educational institutions and scheduling visits on days and times that are more convenient for community members. Moreover, the presence of nearby vaccination centers and hospitals was found to negatively impact MHC utilization. This suggests that MHCs are particularly valuable in areas with limited access to fixed-site vaccination centers, highlighting the importance of strategic placement in underserved regions.

The predictive models identified 1,227 census tracts with higher potential for vaccine uptake, which had a higher rate of uninsurance and mortality, a higher proportion of ethnic minority populations and adolescents, and were in rural areas. These findings are consistent with MHC-centered studies conducted for other states and cities.[36–38] In this study, we found additional healthcare-related factors associated with MHC utility. Census tracts with lower rates of primary care practitioners, hospitals, and other vaccination sites had higher MHC utilization compared to the low-demand census tracts, highlighting the potential of MHC utilization at locations with limited healthcare resources and facilities. The role of predictive models is to act as a classifying or categorization mechanism that identifies areas and communities that are more likely to use the MHCs given the limited data from previous MHC visits. A possible setting for MHC allocation could be to use the existing data and base on the categorical projection of new areas such as census tracts that MHC could possibly get high demand. As MHCs conduct visits to more places relying on the projected high-demand areas, the models can be validated and updated based on the new data. Hence this framework will allow an optimal deployment of MHCs to gain the highest utilization especially by ones who need these services the most.

The time component of projections for MHC utilization at the census tract level provides important decision-making implications for public health planning. Before March 31, 2021, the models predicted high MHC utilization in most census tracts, reflecting the high demand for vaccines during the initial rollout phase. However, after this date, the predicted utilization decreased significantly, indicating the need for continuous assessment and adaptation of MHC deployment strategies as the pandemic evolves and vaccine availability at other providers changes.

This study has several limitations. First, there were substantial changes in vaccine eligibility and availability which affected the MHC utilization. The retrospective design and reliance on limited data may not capture all relevant factors influencing MHC utilization. These factors may also differ from state to state. Future research should consider prospective studies and incorporate additional factors such as community engagement and outreach efforts to provide a more comprehensive understanding of MHC effectiveness. To extend the similar framework to other states, countries, and to different diseases, one needs to consider the determinants of the regions and the disease.

In conclusion, this study provides a framework for optimizing the deployment of MHCs in future health emergencies by identifying factors associated with higher vaccine uptake and predicting areas of high utilization. Predicting the highest- and lowest-demand census tracts for MHC utilization provides categorized importance structure for public health planning and resource allocation. Strategic allocation of MHCs based on these insights can enhance the timely delivery of essential resources to the most vulnerable communities during health emergencies and ultimately save more lives.

## Data Availability

Community level datasets are publicly available and obtained from Centers for Disease Control and Prevention Social Vulnerability Index, the United States Census Bureau American Community Survey and American Hospital Directory.

https://data.census.gov/table/ACSDP5Y2021.DP05?q=acs%202021&g=040XX00US45

https://www.atsdr.cdc.gov/placeandhealth/svi/data_documentation_download.html

https://sc.edu/study/colleges_schools/medicine/centers_and_institutes_new/center_for_rural_and_primary_healthcare/index.php

## Appendix

### Model description

#### Negative binomial generalized linear mixed effects model

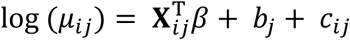

The model assumes the following conditions:

***Y_ij_*** Number of individuals utilized the mobile health clinic (MHC) at i-th site visit in the j-th census tract. The outcome variable *Y_ij_* follows the negative binomial distribution *Y_ij ∼_ NegBin(μ_ij_*, *θ)* where *μ_ij_* is the mean of the negative binomial distribution at i-th site visit in the j-th census tract and θ is the dispersion parameter.

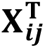: Vector of fixed effects for the i-th site visit in the j-th census tract. Fixed effects included the census tract population, visit term (term 1: before March 31, 2021, and after March 31, 2021), site category (food banks, schools, universities, corporate, homeless shelters, and other), time of the week (Monday to Thursday, Friday, and weekend), time of the week (morning, afternoon, and evening), visit number (first, second, and third or more), and the duration of the visit. The number of vaccination centers and hospitals close to the MHC location is added to the model separately to avoid collinearity between these variables. The visit time changed to (term 1: before May 10, 2021, and after May 10, 2021) for the sensitivity analysis.

***b_j_***: Random effect for the j-th census tract with 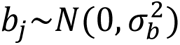

***c_ij_***: Random effect for the i-th site in the j-th census tract 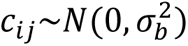

#### Ordinal logistic regression model

Let Y be the ordered categorical outcome of number of individuals utilized the MHC in a census tract {1: 10-19, 2: 20-49, 3: 50-99, 4: 100-249, 5: 250-399, and 6: more than 400 individuals}. Ordinal logistic regression calculates the logged odds of being equal or less than a specific category k of the outcome variable. The cumulative logit model (proportional odds model) can be expressed as:

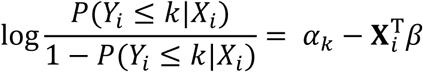

*Y_i_*: The ordered categorical outcome {1: 10-19, 2: 20-49, 3: 50-99, 4: 100-249, 5: 250-399, and 6: more than 400 individuals} of MHC for i-th census tract.

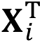: The vector of fixed effects for the i-th census tract. These effects include vaccination term, number of visits conducted to the census tract, census tract level population, proportion of individuals within 30-44, 45-64, and 65+ years of ages, proportion of males, non-White, unemployed, labor force participation, uninsured, under poverty, social vulnerability index (SVI), median income, primary care physicians per 1,000 people, all-cause mortality rate, hospital presence, and percent of rural areas. Variables that are available for zip codes are transformed to census tract level by weighting on the population of a census tract living in different zip codes.

Finally, *α_k_* are the threshold parameters (cut-points) separating the adjacent categories of the ordinal response.

**Table S1:** Characteristics of census tracts that are projected to have high (group: 50-99) and low (groups: 0-19 and 20-49) MHC utilization for COVID-19 vaccination after April 1, 2021. Median values with IQR and p-values for significance of the difference of medians are provided.

|  | High Utilization<br>N = 181 | Low Utilization<br>N = 1,115 | P-value |
| --- | --- | --- | --- |
| % Age under 18 | 21.3 (17.7-25.3) | 21.6 (18.0-25.0) | 0.480 |
| % Age 18-29 | 13.4 (10.5-16.0) | 14.5 (11.4-18.1) | **0.002** |
| % Age 30-44 | 19.3 (15.9-22.6) | 17.7 (14.7-20.9) | **<0.001** |
| % Age 45-64 | 27.3 (23.6-30.6) | 26.3 (22.5-29.7) | **0.006** |
| % Age over 65 | 16.0 (12.7-21.2) | 17.7 (14.1-22.0) | **<0.001** |
| % Male | 48.5 (46.1-50.8) | 48.5 (45.9-51.4) | 0.862 |
| % Non-white | 35.1 (23.0-54.4) | 34.8 (18.9-54.3) | 0.949 |
| % Hispanic | 5.7 (3.6-8.3) | 4.5 (2.6-7.4) | **<0.001** |
| SVI | 0.29 (0.14-0.53) | 0.57 (0.32-0.80) | **<0.001** |
| Income (×\$1000) | 69.8 (55.7-90.1) | 50.5 (39.3-63.2) | **<0.001** |
| % Unemployed | 4.3 (2.2-6.6) | 4.8 (2.7-8.0) | **<0.001** |
| PCP rate | 0.2 (0.0-0.6) | 0.4 (0.1-0.9) | **<0.001** |
| Hospitals | 0.0 (0.0-0.2) | 0.1 (0.0-1.0) | **<0.001** |
| % Uninsured | 9.2 (7.0-11.6) | 11.0 (9.0-12.5) | **<0.001** |
| Mortality rate | 42.3 (33.6-50.1) | 54.0 (44.6-63.2) | **<0.001** |
| % in Poverty | 7.7 (4.8-11.2) | 12.6 (8.8-16.6) | **<0.001** |
| % in Rural | 10.6 (1.8-44.7) | 34.1 (6.8-64.4) | **<0.001** |
| Hospitalizations | 1,271 (796-2,014) | 1,106 (640-1,980) | 0.072 |
| Dead | 57 (32-92) | 64 (33-100) | **0.050** |

**Figure S1:**
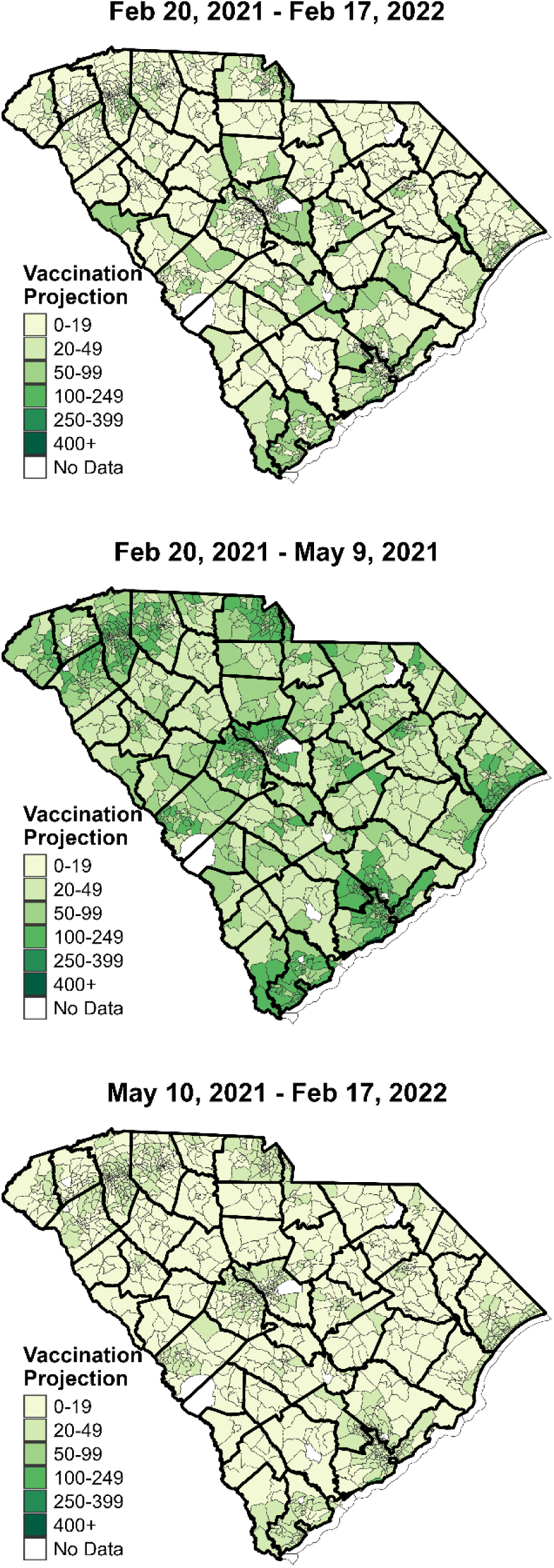
Projected MHC utilization for COVID-19 vaccination at census tracts at different time periods (12-months, and pre and post May 10, 2021).

**Table S2:** Results for prediction accuracy. Number of census tracts that have the same and deviated observed and predicted category for 12-month predictions, and term-based predictions (term 1: before May 9, 2021, term 2: after May 10, 2021).

|  | **12-Month**<br>**Predictions** | **Term-based**<br>**Predictions** |
| --- | --- | --- |
| **Predicted Category** | **N (%)** | **N (%)** |
| 5 groups higher than truth | - | 18 (0.1) |
| 4 groups higher than truth | 4 (0.0) | 11 (0.0) |
| 3 groups higher than truth | 286 (1.1) | 409 (1.6) |
| 2 groups higher than truth | 2,678 (10.7) | 1,841 (7.4) |
| **1 group higher than truth** | **5,110 (20.4)** | **5,898 (23.6)** |
| **Exact group** | **7,158 (28.6)** | **6,539 (26.2)** |
| **1 group lower than truth** | **4,869 (19.5)** | **5,495 (22.0)** |
| 2 group lower than truth | 3,069 (12.3) | 3362 (13.4) |
| 3 group lower than truth | 1,402 (5.6) | 1,213 (4.9) |
| 4 group lower than truth | 365 (1.5) | 214 (0.9) |
| 5 group lower than truth | 47 (0.2) | - |

**Table S3:** Number of census tracts (N) for each category of the projected MHC utilization. Projections are made for an MHC visit at any time during 12-month period (February 20, 2021, to February 17, 2022), before May 9, 2021, and after May 10, 2021.

| **Grouping** | **12-Month**<br>**N = 1,296 (%)** | **Before**<br>**May 9**<br>**N = 1,296 (%)** | **After**<br>**May 10**<br>**N = 1,296 (%)** |
| --- | --- | --- | --- |
| 0-19 | 914 (70.5) | 5 (0.4) | 753 (58.1) |
| 20-49 | 187 (14.4) | 388 (29.9) | 540 (41.7) |
| 50-99 | 192 (14.8) | 455 (35.1) | 2 (0.2) |
| 100-249 | 2 (0.2) | 447 (34.5) | 1 (0.1) |
| 250-399 | 1 (0.1) | - | - |
| 400+ | - | 1 (0.1) | - |

**Table S4:** Characteristics of census tracts that are projected to have high (groups: 50-99, 100- 249, and 400+) and low (groups: 0-19 and 20-49) MHC utilization for COVID-19 vaccination before May 9, 2021. Median values with IQR and p-values for significance of the difference of medians are provided.

|  | **High Utilization**<br>**N = 903** | **Low Utilization**<br>**N = 393** | **P-value** |
| --- | --- | --- | --- |
| % Age under 18 | 21.0 (17.5-24.6) | 22.7 (18.8-26.1) | **<0.001** |
| % Age 18-29 | 13.9 (10.7-17.7) | 14.8 (12.3-18.1) | **0.007** |
| % Age 30-44 | 18.5 (15.2-21.8) | 17.5 (14.2-20.9) | **0.009** |
| % Age 45-64 | 27.1 (23.6-30.5) | 25.1 (21.0-28.6) | **<0.001** |
| % Age over 65 | 17.1 (13.2-22.0) | 17.9 (14.2-21.7) | 0.122 |
| % Male | 48.9 (46.6-51.6) | 47.4 (44.6-50.4) | **<0.001** |
| % Non-white | 31.5 (18.0-51.0) | 39.8 (25.3-59.1) | **<0.001** |
| % Hispanic | 5.4 (3.4-8.5) | 3.1 (2.0-5.8) | **<0.001** |
| SVI | 0.4 (0.2-0.7) | 0.6 (0.4-0.8) | **<0.001** |
| Income (×\$1000) | 61.2 (47.8-78.9) | 43.1 (34.8-53.7) | **<0.001** |
| % Unemployed | 4.5 (2.3-7.0) | 5.1 (3.1-8.4) | **<0.001** |
| PCP rate | 0.3 (0.0-0.7) | 0.5 (0.1-1.5) | **<0.001** |
| Hospitals | 0.0 (0.0-1.0) | 0.2 (0.0-1.0) | **0.001** |
| % Uninsured | 10.7 (8.3-12.6) | 10.7 (9.0-11.8) | 0.727 |
| Mortality rate | 48.0 (38.6-56.8) | 58.7 (49.6-66.1) | **<0.001** |
| % in Poverty | 10.1 (6.1-14.5) | 14.4 (10.6-18.4) | **<0.001** |
| % in Rural | 14.9 (2.4-45.7) | 59.7 (33.2-92.5) | **<0.001** |
| COVID-19 ED visits | 1,339 (796-2,091) | 843 (430-1,432) | **<0.001** |
| COVID-19 deaths | 67 (40-104) | 46 (22-76) | **<0.001** |

**Table S5:** Characteristics of census tracts that are projected to have high (groups: 20-49, 50-99, and 100-249) and low (group: 0-19) MHC utilization for COVID-19 vaccination after May 10, 2021. Median values with IQR and p-values for significance of the difference of medians are provided.

|  | **High Utilization**<br>**N = 543** | **Low Utilization**<br>**N = 753** | **P-value** |
| --- | --- | --- | --- |
| % Age under 18 | 20.8 (17.4-24.4) | 22.0 (18.1-25.5) | **0.001** |
| % Age 18-29 | 13.2 (10.0-16.8) | 14.8 (12.2-18.5) | **<0.001** |
| % Age 30-44 | 18.5 (15.1-21.8) | 18.0 (14.8-21.2) | 0.132 |
| % Age 45-64 | 27.7 (24.5-31.2) | 25.7 (21.8-29.1) | **<0.001** |
| % Age over 65 | 16.7 (13.0-22.4) | 17.7 (14.0-21.7) | **0.013** |
| % Male | 49.0 (47.0-51.5) | 48.1 (45.0-51.0) | **<0.001** |
| % Non-white | 28.4 (16.1-46.4) | 39.3 (23.9-58.4) | **<0.001** |
| % Hispanic | 5.9 (3.8-9.4) | 4.0 (2.3-6.7) | **<0.001** |
| SVI | 0.3 (0.1-0.6) | 0.6 (0.4-0.8) | **<0.001** |
| Income (×\$1000) | 68.6 (55.9-88.0) | 46.6 (37.1-56.9) | **<0.001** |
| % Unemployed | 4.1 (2.2-6.2) | 5.2 (3.0-8.3) | **<0.001** |
| PCP rate | 0.2 (0.0-0.6) | 0.4 (0.1-1.1) | **<0.001** |
| Hospitals | 0.0 (0.0-1.0) | 0.1 (0.0-1.0) | **<0.001** |
| % Uninsured | 10.4 (7.9-12.6) | 10.8 (9.0-12.1) | 0.063 |
| Mortality rate | 43.7 (36.0-53.0) | 56.0 (47.4-64.2) | **<0.001** |
| % in Poverty | 8.2 (5.3-12.1) | 13.5 (9.7-17.3) | **<0.001** |
| % in Rural | 7.6 (1.4-28.1) | 51.2 (17.8-82.6) | **<0.001** |
| COVID-19 ED visits | 1,478 (904-2,349) | 953 (515-1,635) | **<0.001** |
| COVID-19 deaths | 69 (43-104) | 56 (26-90) | **<0.001** |

**Table S6:** Characteristics of census tracts that are projected to have high (groups: 20-49, 50-99, 100-249, and 250-399) and low (group: 0-19) MHC utilization for COVID-19 vaccination any time during 12-month period. Median values with IQR and p-values for significance of the difference of medians are provided.

|  | High Utilization<br>N = 382 | Low Utilization<br>N = 914 | P-value |
| --- | --- | --- | --- |
| % Age under 18 | 21.3 (17.3-25.5) | 21.6 (18.1-24.9) | 0.472 |
| % Age 18-29 | 13.1 (9.9-16.0) | 14.6 (11.9-18.4) | **<0.001** |
| % Age 30-44 | 18.9 (14.9-22.4) | 17.9 (14.9-21.0) | **0.008** |
| % Age 45-64 | 26.8 (23.1-30.1) | 26.4 (22.7-29.8) | 0.267 |
| % Age over 65 | 17.3 (13.3-22.9) | 17.3 (13.5-21.4) | 0.894 |
| % Male | 48.7 (46.3-51.2) | 48.4 (45.8-51.2) | 0.190 |
| % Non-white | 35.7 (22.2-57.4) | 34.6 (19.6-52.0) | 0.587 |
| % Hispanic | 5.2 (3.1-8.1) | 4.6 (2.7-7.5) | **0.043** |
| SVI | 0.4 (0.1-0.7) | 0.6 (0.3-0.8) | **<0.001** |
| Income (×\$1000) | 69.6 (53.2-91.3) | 50.4 (39.2-62.6) | **<0.001** |
| % Unemployed | 4.5 (2.3-7.1) | 4.7 (2.7-7.6) | **<0.001** |
| PCP rate | 0.2 (0.0-0.6) | 0.4 (0.1-1.0) | **<0.001** |
| Hospitals | 0.0 (0.0-0.4) | 0.2 (0.0-1.0) | **<0.001** |
| % Uninsured | 9.6 (7.3-12.0) | 10.9 (9.0-12.4) | **<0.001** |
| Mortality rate | 44.6 (35.3-53.9) | 53.6 (44.6-62.6) | **<0.001** |
| % in Poverty | 8.1 (4.8-11.8) | 12.7 (9.2-16.5) | **<0.001** |
| % in Rural | 15.0 (1.8-49.3) | 34.0 (6.9-64.3) | **<0.001** |
| COVID-19 ED visits | 1,155 (627-1,999) | 1,151 (661-2,005) | 0.951 |
| COVID-19 deaths | 54 (29-87) | 66 (36-101) | **<0.001** |

## References

1. CDC. Centers for Disease Control and Prevention. 2020 [cited 2023 Sep 14]. COVID Data Tracker. Available from: https://covid.cdc.gov/covid-data-tracker

2. Risk for COVID-19 infection, hospitalization, and death by race/ethnicity. [cited 2024 Jun 28]; Available from: https://stacks.cdc.gov/view/cdc/105453

3. Henning-Smith C. The Unique Impact of COVID-19 on Older Adults in Rural Areas. J Aging Soc Policy. 2020;32(4–5):396–402.

4. Paul R, Arif A, Pokhrel K, Ghosh S. The Association of Social Determinants of Health With COVID-19 Mortality in Rural and Urban Counties. J Rural Health. 2021;37(2):278–86.

5. Overcoming Barriers to COVID-19 Vaccination in African Americans: The Need for Cultural Humility [Internet]. [cited 2024 Jun 28]. Available from: https://ajph.aphapublications.org/doi/epub/10.2105/AJPH.2020.306135

6. Lieberman-Cribbin W, Tuminello S, Flores RM, Taioli E. Disparities in COVID-19 Testing and Positivity in New York City. Am J Prev Med. 2020 Sep;59(3):326–32.

7. Press VG, Huisingh-Scheetz M, Arora VM. Inequities in Technology Contribute to Disparities in COVID-19 Vaccine Distribution. JAMA Health Forum. 2021 Mar 19;2(3):e210264.

8. Fisk RJ. Barriers to vaccination for coronavirus disease 2019 (COVID-19) control: experience from the United States. Glob Health J. 2021 Mar 1;5(1):51–5.

9. Abdul-Mutakabbir JC, Casey S, Jews V, King A, Simmons K, Hogue MD, et al. A three-tiered approach to address barriers to COVID-19 vaccine delivery in the Black community. Lancet Glob Health. 2021 Jun;9(6):e749–50.

10. Rutter H, Savona N, Glonti K, Bibby J, Cummins S, Finegood DT, et al. The need for a complex systems model of evidence for public health. The Lancet. 2017 Dec 9;390(10112):2602–4.

11. Yu SWY, Hill C, Ricks ML, Bennet J, Oriol NE. The scope and impact of mobile health clinics in the United States: a literature review. Int J Equity Health. 2017 Oct 5;16(1):178.

12. Schwitters A, Lederer P, Zilversmit L, Gudo PS, Ramiro I, Cumba L, et al. Barriers to health care in rural Mozambique: a rapid ethnographic assessment of planned mobile health clinics for ART. Glob Health Sci Pract. 2015 Mar;3(1):109–16.

13. Attipoe-Dorcoo S, Delgado R, Gupta A, Bennet J, Oriol NE, Jain SH. Mobile health clinic model in the COVID-19 pandemic: lessons learned and opportunities for policy changes and innovation. Int J Equity Health. 2020 May 19;19(1):73.

14. Rassekh BM, Shu W, Santosham M, Burnham G, Doocy S. An evaluation of public, private, and mobile health clinic usage for children under age 5 in Aceh after the tsunami: implications for future disasters. Health Psychol Behav Med. 2014 Jan 1;2(1):359–78.

15. Larson R, Hussain S, Chau MM, Jones A, Vangeepuram N, Madden D, et al. The Power of Partnership: NYCEAL Collaborations With Health Agencies and Mobile Vaccination Vans. Am J Public Health. 2024 Jan;114(Suppl 1):S92–5.

16. Kulle AC, Schumacher S, von Bieberstein F. Mobile vaccination units substantially increase COVID-19 vaccinations: evidence from a randomized controlled trial. J Public Health. 2024 Mar 1;46(1):151–7.

17. Levy P, McGlynn E, Hill AB, Zhang L, Korzeniewski SJ, Foster B, et al. From pandemic response to portable population health: A formative evaluation of the Detroit mobile health unit program. PLoS ONE. 2021 Nov 30;16(11):e0256908.

18. Zhang X, Tulloch JSP, Knott S, Allison R, Parvulescu P, Buchan IE, et al. Evaluating the impact of using mobile vaccination units to increase COVID-19 vaccination uptake in Cheshire and Merseyside, UK: a synthetic control analysis. BMJ Open. 2023 Oct 6;13(10):e071852.

19. Serrano-Alarcón M, Mckee M, Palumbo L, Salvi C, Johansen A, Stuckler D. How to increase COVID-19 vaccination among a population with persistently suboptimal vaccine uptake? Evidence from the North Macedonia mobile vaccination and public health advice caravan. Health Policy Amst Neth. 2024 Jan;139:104966.

20. Alvi RA, Justason L, Liotta C, Martinez-Helfman S, Dennis K, Croker SP, et al. The Eagles Eye Mobile: assessing its ability to deliver eye care in a high-risk community. J Pediatr Ophthalmol Strabismus. 2015;52(2):98–105.

21. Hill C, Zurakowski D, Bennet J, Walker-White R, Osman JL, Quarles A, et al. Knowledgeable Neighbors:A Mobile Clinic Model for Disease Prevention and Screening in Underserved Communities. Am J Public Health. 2012 Mar;102(3):406–10.

22. Emanuel EJ, Persad G, Upshur R, Thome B, Parker M, Glickman A, et al. Fair Allocation of Scarce Medical Resources in the Time of Covid-19. N Engl J Med. 2020 May 21;382(21):2049–55.

23. Woolf SH, Chapman DA, Lee JH. COVID-19 as the Leading Cause of Death in the United States. JAMA [Internet]. 2020 Dec 17 [cited 2021 Mar 29]; Available from: https://jamanetwork.com/journals/jama/fullarticle/2774465

24. The New York Times. Coronavirus in the U.S.: Latest Map and Case Count. The New York Times [Internet]. 2020 Mar 3 [cited 2022 Feb 28]; Available from: https://www.nytimes.com/interactive/2021/us/covid-cases.html

25. Brown KA, Stall NM, Joh E, Allen U, Bogoch II, Buchan SA, et al. COVID-19 Vaccination Strategy for Ontario Using Age and Neighbourhood-Based Prioritization. Sci Briefs Ont COVID-19 Sci Advis Table. 2021 Feb 26;2(10).

26. Salmon D, Opel DJ, Dudley MZ, Brewer J, Breiman R. Reflections On Governance, Communication, And Equity: Challenges And Opportunities In COVID-19 Vaccination. Health Aff (Millwood). 2021 Feb 4;40(3):419–25.

27. Bachireddy C, Dar M, Chen C. Medicaid and COVID-19 Vaccination—Translating Equitable Allocation Into Equitable Administration. JAMA Health Forum. 2021 Feb 17;2(2):e210114.

28. Rutter H, Savona N, Glonti K, Bibby J, Cummins S, Finegood DT, et al. The need for a complex systems model of evidence for public health. The Lancet. 2017 Dec 9;390(10112):2602–4.

29. Quinn SC, Kumar S. Health Inequalities and Infectious Disease Epidemics: A Challenge for Global Health Security. Biosecurity Bioterrorism Biodefense Strategy Pract Sci. 2014 Sep 1;12(5):263–73.

30. Guttieres D, Sinskey AJ, Springs SL. Modeling Framework to Evaluate Vaccine Strategies against the COVID-19 Pandemic. Systems. 2021 Mar;9(1):4.

31. Angulo FJ, Finelli L, Swerdlow DL. Reopening Society and the Need for Real-Time Assessment of COVID-19 at the Community Level. JAMA. 2020;323(22):2247–8.

32. Rennert L, McMahan C, Kalbaugh CA, Yang Y, Lumsden B, Dean D, et al. Surveillance-based informative testing for detection and containment of SARS-CoV-2 outbreaks on a public university campus: an observational and modelling study. Lancet Child Adolesc Health. 2021;5(6):428–36.

33. Rennert L, Howard KA, Kickham CM, Gezer F, Coleman A, Roth P, et al. Implementation of a mobile health clinic framework for Hepatitis C virus screening and treatment: a descriptive study. Lancet Reg Health – Am [Internet]. 2024 Jan 1 [cited 2024 Apr 2];29. Available from: https://www.thelancet.com/journals/lanam/article/PIIS2667-193X(23)00222-3/fulltext

34. Rennert L, Gezer F, Jayawardena I, Howard KA, Bennett KJ, Litwin AH, et al. Mobile health clinics for distribution of vaccinations to underserved communities during health emergencies: A COVID-19 case study. Public Health Pract. 2024;Under Review.

35. Gezer F, Howard KA, Litwin AH, Martin NK, Rennert L. Identification of factors associated with opioid-related and hepatitis C virus-related hospitalisations at the ZIP code area level in the USA: an ecological and modelling study. Lancet Public Health. 2024 Jun 1;9(6):e354–64.

36. Alcendor DJ, Juarez PD, Matthews-Juarez P, Simon S, Nash C, Lewis K, et al. Meharry Medical College Mobile Vaccination Program: Implications for Increasing COVID-19 Vaccine Uptake among Minority Communities in Middle Tennessee. Vaccines. 2022 Jan 29;10(2):211.

37. Gupta PS, Mohareb AM, Valdes C, Price C, Jollife M, Regis C, et al. Expanding COVID-19 vaccine access to underserved populations through implementation of mobile vaccination units. Prev Med. 2022 Oct 1;163:107226.

38. Gavin RM, Countryman M, Musco J, Ricard R, Roberts A, Lees C. Reaching Diverse Communities During a Local Public Health COVID-19 Vaccination Response Through a Mobile Clinic Compared to Mass Vaccination Sites. J Public Health Manag Pract. 2024 Jun;30(3):411.

39. Schmitz M, Luminet O, Klein O, Morbée S, Van den Bergh O, Van Oost P, et al. Predicting vaccine uptake during COVID-19 crisis: A motivational approach. Vaccine. 2022 Jan 21;40(2):288–97.

40. Cheong Q, Au-yeung M, Quon S, Concepcion K, Kong JD. Predictive Modeling of Vaccination Uptake in US Counties: A Machine Learning–Based Approach. J Med Internet Res. 2021 Nov 25;23(11):e33231.

41. Shiloh S, Peleg S, Nudelman G. Vaccination Against COVID-19: A Longitudinal Trans- Theoretical Study to Determine Factors that Predict Intentions and Behavior. Ann Behav Med. 2022 Apr 1;56(4):357–67.

42. Zhou X, Li Y. Forecasting the COVID-19 vaccine uptake rate: an infodemiological study in the US. Hum Vaccines Immunother. 2022 Jan 31;18(1):2017216.

43. Prisma Health celebrates groundbreaking of new emergency department at Prisma Health Oconee Memorial Hospital [Internet]. [cited 2024 Aug 13]. Available from: https://prismahealth.org/patients-and-guests/news/prisma-health-celebrates-groundbreaking-of-new-emergency-department-at-prisma-health-oconee-memorial

44. DP05: ACS DEMOGRAPHIC AND … - Census Bureau Table [Internet]. [cited 2024 Sep 5]. Available from: https://data.census.gov/table/ACSDP5Y2021.DP05?q=acs%202021&g=040XX00US45

45. CDC/ATSDR SVI Data and Documentation Download | Place and Health | ATSDR [Internet]. 2022 [cited 2023 Mar 7]. Available from: https://www.atsdr.cdc.gov/placeandhealth/svi/data_documentation_download.html

46. Flanagan BE, Gregory EW, Hallisey EJ, Heitgerd JL, Lewis B. A Social Vulnerability Index for Disaster Management. J Homel Secur Emerg Manag [Internet]. 2011 Jan 5 [cited 2023 Mar 7];8(1). Available from: https://www.degruyter.com/document/doi/10.2202/1547-7355.1792/html

47. South Carolina Center for Rural and Primary Healthcare - School of Medicine Columbia | University of South Carolina [Internet]. [cited 2024 Apr 2]. Available from: https://sc.edu/study/colleges_schools/medicine/centers_and_institutes_new/center_for_rural_and_primary_healthcare/index.php

48. COVID-19 Vaccines Providers [Internet]. [cited 2024 Jul 8]. Available from: https://sc-dhec.maps.arcgis.com/apps/instant/nearby/index.html?appid=903e4c38fc4b4771b04fb492d366fa78

49. American Hospital Directory - Free Daily Limit [Internet]. [cited 2024 Apr 8]. Available from: https://www.ahd.com/states/hospital_SC.html

50. Healthcare | South Carolina Revenue and Fiscal Affairs Office [Internet]. [cited 2023 Mar 7]. Available from: https://rfa.sc.gov/data-research/healthcare

51. All South Carolinians Aged 16 and Older to be Eligible for COVID-19 Vaccine Beginning March 31, 2021 | S.C. Governor Henry McMaster [Internet]. [cited 2024 Jul 8]. Available from: https://governor.sc.gov/news/2021-03/all-south-carolinians-aged-16-and-older-be-eligible-covid-19-vaccine-beginning-march

52. AJMC [Internet]. 2021 [cited 2023 Sep 5]. A Timeline of COVID-19 Vaccine Developments in 2021. Available from: https://www.ajmc.com/view/a-timeline-of-covid-19-vaccine-developments-in-2021

